# Single-cell characterization of peripheral blood mononuclear cells from vedolizumab-treated patients with Crohn’s disease identifies response-associated differences among the plasmacytoid dendritic cell and classical monocyte populations

**DOI:** 10.1101/2023.06.23.23291732

**Authors:** Andrew Y.F. Li Yim, Ishtu L. Hageman, Vincent Joustra, Ahmed Elfiky, Mohammed Ghiboub, Evgeni Levin, Jan Verhoeff, Caroline Verseijden, Iris Admiraal-van den Berg, Marcel M.A.M. Mannens, Marja E. Jakobs, Susan B. Kenter, Alex T. Adams, Jack Satsangi, Geert R. D’Haens, Wouter J. de Jonge, Peter Henneman

## Abstract

Vedolizumab (VDZ) is a monoclonal antibody approved for the treatment of Crohn’s disease (CD). Despite its efficacy, non-response to VDZ is common in clinical practice with no clear understanding of how it manifests. Here, we characterized the cellular repertoire of responders and non-responders to VDZ during treatment. Peripheral blood mononuclear cells (PBMCs) were isolated from CD patients on VDZ treatment that were either steroid-free responder (N = 4) or non-responder (N = 4). Response was defined as ≥3 drop in Simple Endoscopic Score for Crohn’s Disease (SES-CD) in combination with a ≥50% reduction in C-reactive protein (CRP) and fecal calprotectin and/or a ≥3 point drop in Harvey-Bradshaw Index (HBI). Single-cell repertoires were characterized using single-cell RNA-sequencing (scRNAseq) and mass cytometry by time of flight (CyTOF). Non-responders to VDZ presented more T cells, but fewer myeloid cells. T cells from non-responders presented lower expression of NFкB signaling inhibitors. A lower relative abundance of plasmacytoid dendritic cells (pDCs) was observed among non-responders. Moreover, non-responder-derived classical monocytes presented lower expression of genes involved in wound-healing and cytokine-cytokine receptor signaling. Taken together, non-response to VDZ during treatment is associated with differences in abundance and expression among T and myeloid cells.

## Introduction

Crohn’s disease (CD) is an incurable, chronic, inflammatory condition of the gastrointestinal tract characterized by a relapsing-remitting transmural inflammation of the digestive tract belonging to the family of inflammatory bowel diseases (IBD). Current treatments for CD include the use of monoclonal antibodies that target mediators of inflammation with the goal of ameliorating the inflammatory phenotype and/or maintaining a state of clinical and endoscopic remission. One such monoclonal antibody is vedolizumab (VDZ), which was approved for use in patients with CD in 2014 by the United States Food and Drug Administration as well as the European Medicines Agency (3).

VDZ targets the gut homing receptor complex integrin α_4_β_7_ (also known as lymphocyte Peyer’s patch adhesion molecule 1; LPAM-1) (4, 5), which prevents it from binding mucosal vascular addressin cell adhesion molecule 1 (MAdCAM-1), a molecule expressed exclusively by the intestinal endothelial cells. By preventing integrin α_4_β_7_ from binding MAdCAM-1, the attachment and stabilization of circulating immune cells that express integrin α_4_β_7_ to high endothelial venules in the gut is destabilized, thereby abrogating gut-homing capabilities (6–8). While VDZ has traditionally been discussed within the context of the T cell lineage (9–12), more recent studies suggest that the myeloid (13, 14) as well as B cells (15) are affected by VDZ treatment as well. Despite the advances VDZ therapy has provided patient care, the efficacy or therapy response rate is reported to be approximately between 30% to 45% (3, 16–18) with a recent meta-analysis indicating that loss of response towards VDZ among CD patients was estimated at 47.9 per 100 person-years (19). To date, we have no proper understanding why only a subgroup of patient responds to therapy, nor do we have a prognostic biomarker for predicting response to VDZ therapy. To better understand how response to VDZ manifests, we conducted an exploratory case-control study to characterize the immune cell composition of peripheral blood mononuclear cells (PBMCs) from CD patients on VDZ treatment. Specifically, we compared responders with non-responders using single-cell RNA-sequencing (scRNAseq) and cytometry by time of flight (CyTOF).

## Results

### Cohort assembly

Peripheral blood samples were obtained from a cohort of patients with CD on VDZ treatment at the AmsterdamUMC, location AMC as part of routine care. Response to treatment was defined as endoscopic-(≥50% drop in simple endoscopic score for Crohn’s Disease (SES-CD)) in combination with biochemical (≥50% reduction in C-reactive protein (CRP) and fecal calprotectin (FCP) or an absolute CRP<5.0 µg/g and FCP<250 µg/g) and/or clinical response (≥3 point drop in Harvey-Bradshaw Index (HBI)) compared to the start of treatment. All patients presented with measurable drug serum concentrations and no concomitant corticosteroid usage. For this study, we selected a cohort of 8 CD patients at a median of 41 weeks into VDZ treatment, which were classified as responder (N = 4) and non-responder (N = 4) (**Methods**, **Table 1**, and **Figure 1**).

**Figure 1.**
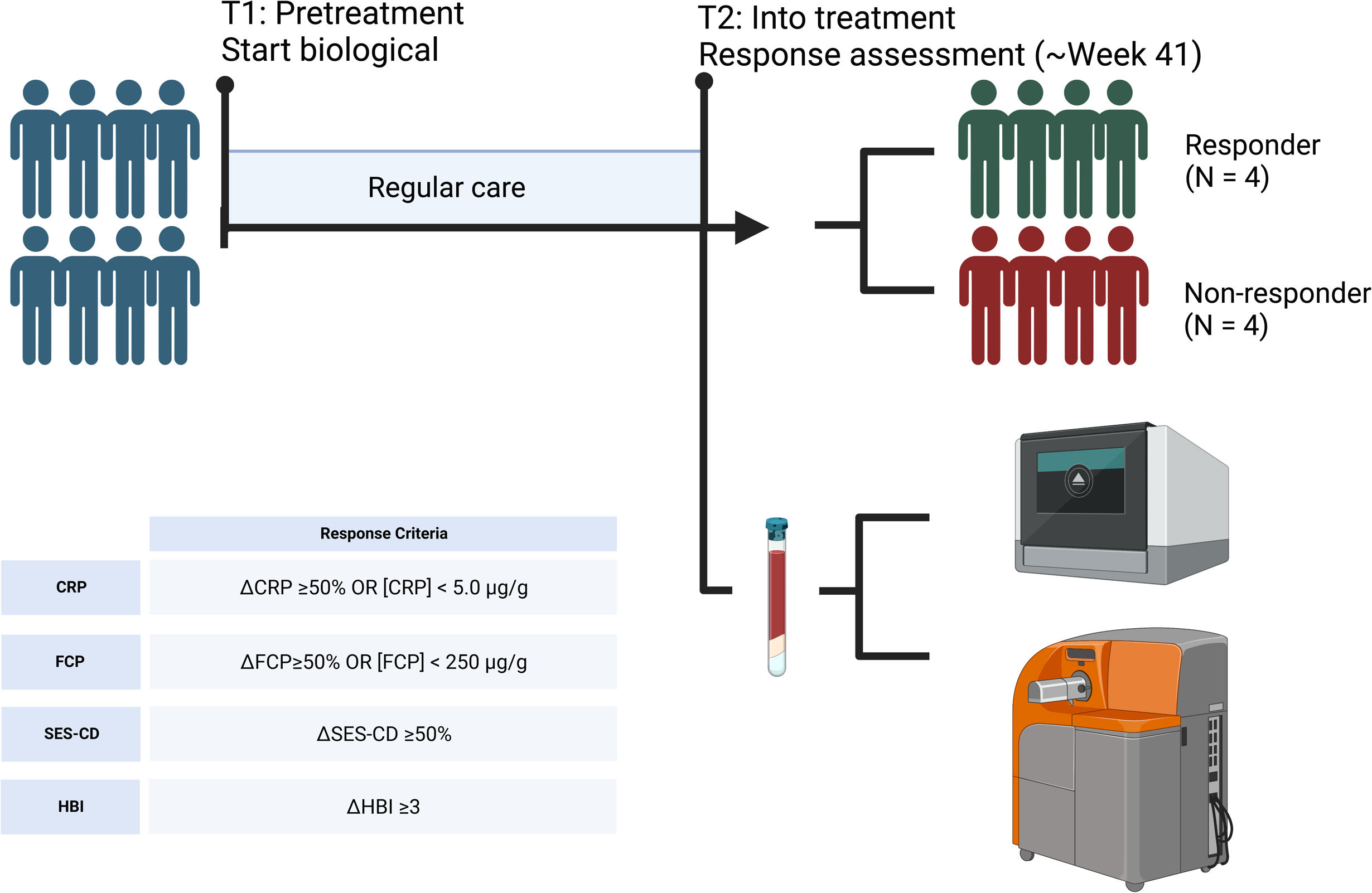
Sampling strategy. Peripheral blood samples were obtained from 4 CD patients that responded to VDZ and 4 CD patients that did not respond to VDZ. Response was defined based the Harvey Bradshaw Index (HBI), C-reactive protein (CRP), fecal calprotectin (FCP), and simple-endoscopic score CD (SES-CD). From peripheral blood, peripheral blood mononuclear cells (PBMCs) were isolated which were subsequently used for single-cell RNA-sequencing and mass cytometry by time of flight (CyTOF) using the Chromium controller (10X Genomics) and CyTOF3-Helios systems (Fluidigm), respectively. Created with BioRender.

**Table 1.** Patient characteristics at time of sampling included in the scRNAseq and CyTOF. Overview of the demographics of the included patients. IQR: Interquartile range. T1: Start of treatment. T2: Response assessment. SES-CD: Simple endoscopic score for Crohn’s disease. HBI: Harvey bradshaw index. CRP: C-reactive protein. FCP: Fecal calprotectin. ADA: Adalimumab. IFX: Infliximab.

|  | Responders (N = 4) | Non-responders (N = 4) |
| --- | --- | --- |
| Female, N (%) | 3 (75) | 4 (100) |
| Age, years, median (IQR) | 40 (35-48) | 48 (36.5-61.25) |
| Disease duration, years, median | 10.5 (7.5-18.5) | 7 (4.5-23) |
| (IQR) |  |  |
| Ethnic background, N (%) |  |  |
| - Caucasian | 3 (75) | 1 (25) |
| SES-CD, median (IQR) |  |  |
| - T1 | 5.5 (5-8) | 6 (6-6.5) |
| - T2 | 1.5 (0-4) | 3 (1.5-3.5) |
| HBI, median (IQR) |  |  |
| - T1 | 9.5 (6-14.5) | 8 (8-9) |
| - T2 | 2.5 (1-4.3) | 9 (6.5-11.3) |
| CRP, mg/L, median (IQR) |  |  |
| - T1 | 9.8 (6.5-14.4) | 2.2 (1.4-2.8) |
| - T2 | 7.6 (5.5-8.9) | 3.1 (0.8-5.9) |
| FCP, µg/g, median (IQR) |  |  |
| - T1 | 328.5 (168-849.5) | 262.5 (189-305.3) |
| - T2 | 54.5 (24.8-168) | 442 (250.5-522) |
| Disease location, n (%) |  |  |
| - Ileal disease (L1) | 3 (75) | 1 (25) |
| - Colonic disease (L2) | - | - |
| - Ileocolonic disease (L3) | 1 (25) | 3 (75) |
| Disease behavior, N (%) |  |  |
| - Non structuring/penetrating (B1) | 1 (25) | 2 (50) |
| - Stricturing (B2) | 2 (50) | 2 (50) |
| - Penetrating (B3) | 1 (25) | - |
| - Perianal disease (p) | 1 (25) | - |
| Previous IBD-related surgery, N (%) | 2 (50) | 1 (25) |
| Concomitant medication, N (%) |  |  |
| - Immunomodulators | - | - |
| - Prednisone | - | - |
| Previous treatment exposure, N (%) |  |  |
| - Immunomodulators | 3 (75) | 1 (25) |
| - Anti-TNF (ADA or IFX) | 1 (25) | 3 (75) |
| Smoking, N (%) |  |  |
| - Never | 1 (25) | 1 (25) |
| - Active | - | 3 (75) |
| - Former | 3 (75) | - |

### ITGA4 expression detected on all PBMCs

Single-cell RNA-sequencing (scRNAseq) and mass cytometry by time of flight (CyTOF) provided transcriptional and proteomic profiles of 15,981 and 1,783,641 (subsampled to 16,000 for comparability purposes) cells, respectively. Cells were annotated to 31 known cell types (**Figure 2A-B**) using a combination of automatic and manual curation based on canonical markers (**Figure 2C-D**). We observed general agreement between the two experiments (r = 0.73; **Figure 2E**). Interrogation of the of the protein expression of integrin α_4_ as well as its encoding gene *ITGA4* were measurably expressed in all cell types (**Figure 2F-G**). By contrast, gene expression of *ITGB7* was notably muted, with the plasma cells presenting the highest expression. Nonetheless, our observations confirm that genes encoding integrin α_4_β_7_ expression are active in many cell types and are hence not solely restricted to the T cells.

**Figure 2.**
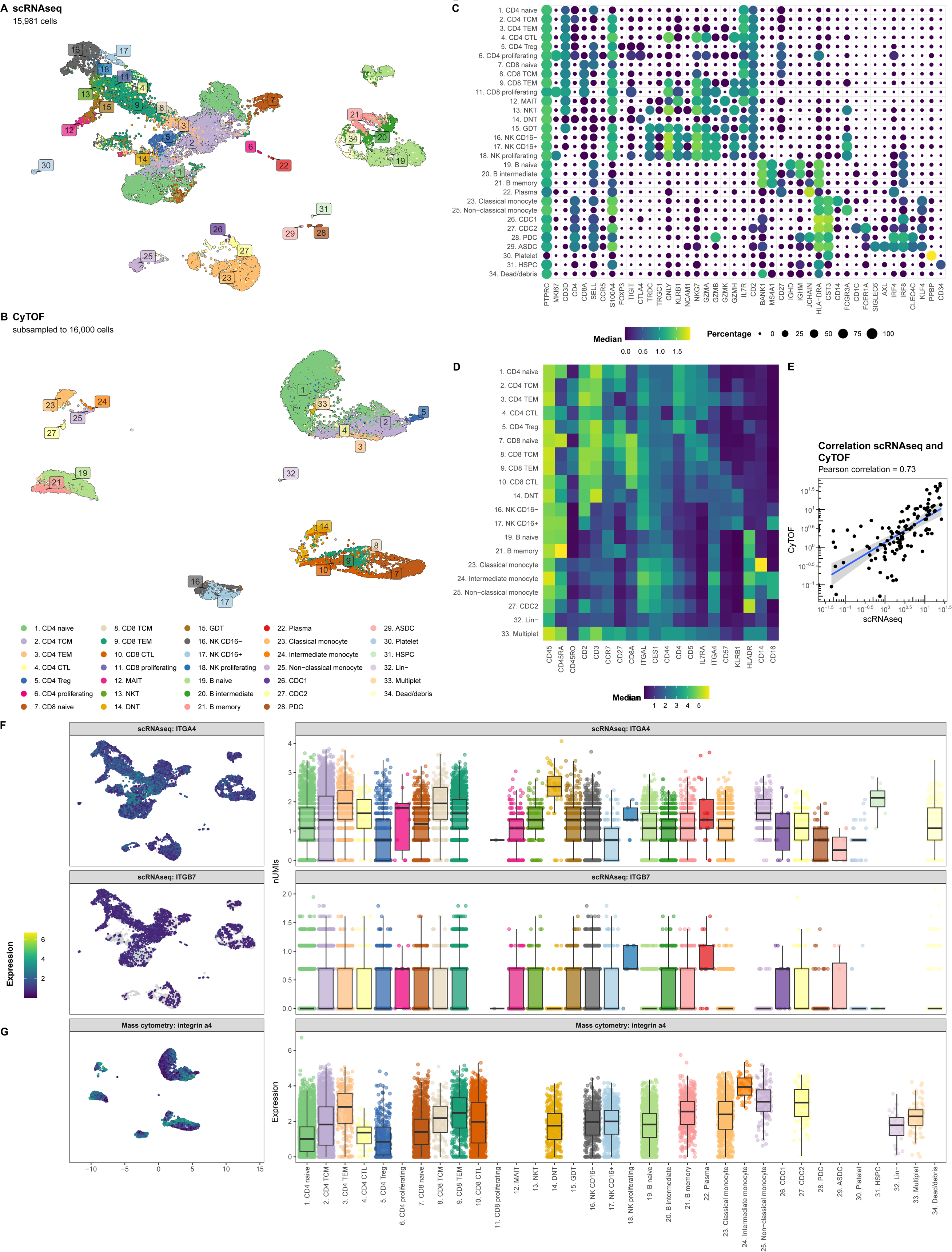
*ITGA4* is expressed by all cell types. Uniform manifold approximation and projection (UMAP) visualization of the PBMCs from CD patients on VDZ that respond (R; N = 4) and that do not respond (NR; N = 4) colored by the cellular identity as obtained through (A) single-cell RNA-sequencing (scRNAseq) and (B) mass cytometry by time of flight (CyTOF). Visualization of the marker expression used to annotate the PBMCs at the level of (C) gene expression through a dotplot where size and color intensity represent the percentage cells with measurable expression and the median expression, respectively, and (D) protein expression through a heatmap with the color representing the median expression. (E) Scatterplot representing the percentage cell types per sample relative to all PBMCs for scRNAseq on the X-axis and CyTOF on the Y-axis colored by lineage show general agreement between the scRNAseq and CyTOF experiment. UMAP (left) and boxplot (right) visualization of the gene expression for (F) *ITGA4*, *ITGB7*, as well as (G) the protein expression for integrin α_4_ per cell type. Colors in the UMAP visualization represent the level of expression per cell where grey represents no measurable expression.

### Circulating T cells from VDZ non-responders express lower levels of inhibitors of the NFкB signaling pathway

Differential abundance analysis of both the scRNAseq and CyTOF data indicated concordant differences between responders and non-responders (**Figure 3A-C** and **Supplemental Table 3**). Overall, non-responders presented with a significantly lower relative abundance of the myeloid (*p*-value = 4.6E-03) cells, whereas the T cells presented a higher abundance (*p*-value = 0.029) (**Figure 3D**). At a more granular level, a significantly higher abundance of CD8 T central memory (CD8 TCM) was observed through CyTOF (*p*-value = 0.01), which we could reproduce in direction, but not in significance, through scRNAseq (*p*-value = 0.89) (**Figure 3E**). At scRNAseq level, a significantly higher abundance was observed for the mucosal associated invariant T cells (MAIT; *p*-value = 0.029) (**Figure 3F**) and a lower abundance of plasmacytoid dendritic cells (pDCs; *p*-value = 0.041) (**Figure 3G**), respectively, which we were unable to reproduce using CyTOF as no markers were included for either MAIT or pDCs. As VDZ reportedly binds T cells in particular (9), we investigated whether their transcriptome presented response-associated differences (**Supplemental Table 4**). We specifically interrogated *ITGA4* and *ITGB7* expression but found no differences in expression for *ITGB7*. By contrast, *ITGA4* was found to be significantly higher in non-responders when looking at CD4 TEM, CD4 Treg, CD8 TCM, and CD8 TEM (**Figure 3H)**. Notably, we observed for multiple T cell subsets that genes encoding inhibitors of the NFкB signaling pathway, such as *TNFAIP3* and *NFKBIA*, were significantly lower in expression amongst non-responders (**Figure 3H**).

**Figure 3.**
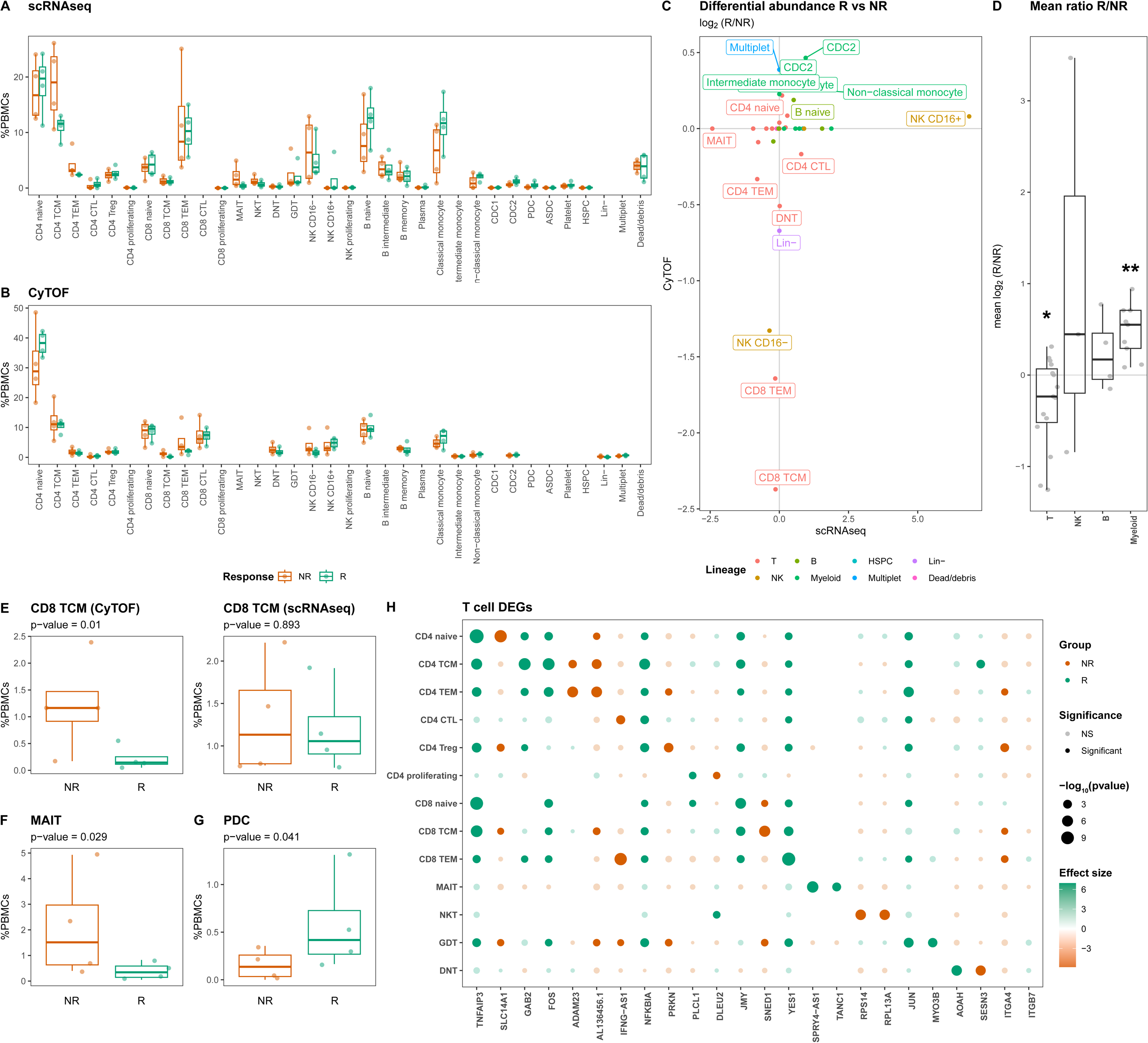
T cells present response-associated differences in abundance and expression. Boxplot visualizations of the cell type abundances relative to all measured PBMCs colored by response for (A) scRNAseq and (B) CyTOF. (C) Scatterplot comparing the differences in abundance based on scRNAseq on the X-axis and CyTOF on the Y-axis. Values represent log_2_-transformed responder:non-responder ratios per cell type and colors represent the parent lineages of each cell type. (D) Boxplot visualizations of the mean scRNAseq-CyTOF log_2_-transformed responder:non-responder ratios shows that T and myeloid cells are significantly more and less abundant amongst non-responders, respectively. Asterisks denote statistical significance using a one-sample t-test against 0. * p<0.05; ** p<0.01. Boxplot visualizations of the abundance (E) CD8 T central memory (CD8 TCM) in the (left) CyTOF experiment and (right) scRNAseq experiment, (F) mucosal associated invariant T (MAIT) cells, and (G) plasmacytoid dendritic cells (pDC) relative to all PBMCs. *P*-values were calculated using the t-test implementation in speckle::propeller. (H) Dotplot visualization representing the most significant differentially expressed genes, as well as *ITGA4* and *ITGB7* per T cell subset. Size represents statistical significance, transparency the significance threshold, and color whether the gene is upregulated in either responders (green) or non-responders (orange). A notable lower expression of *TNFAIP3* and *NFKBIA* can be observed among non-responders.

### VDZ non-responders present higher relative abundances of circulating plasmacytoid dendritic cells

As our CyTOF panel did not include markers for pDCs, we conducted flow cytometry analyses where we identified the (T/B/NK)Lin^-^HLA-DR^+^CD11c^-^CD123^+^ pDC fraction (**Figure 4A**). Indeed, we observed a significantly lower proportion of circulatory pDCs among responders relative to the non-responders (*p*-value = 0.03) (**Figure 4B**). However, interrogating the transcriptome of the pDCs did not identify statistically significant response-associated differences between responders and non-responders after correcting for multiple testing (**Figure 4C** and **Supplemental Table 5**). Notably, the expression of *ITGA4* and *ITGB7* specifically indicated a lower expression of *ITGB7* amongst non-responders albeit statistically non-significant (*p*-value = 0.32) (**Figure 4D** and **Supplemental Table 5**). We hypothesized that the diminished proportion of circulatory pDCs among non-responders was due to their recruitment into the gastrointestinal tract thereby removing them from circulation. To corroborate our hypothesis, we interrogated the publicly available single-cell transcriptomic data from CD patients’ intestinal biopsies extracted from ileal lesions (involved) and adjacent non-lesional (uninvolved) tissue as published by Martin *et al.* (20). Upon identifying the pDC fraction (**Figure 4E**), we found that the pDC proportion relative to the total immune fraction was suggestively higher in lesional compared to non-lesional areas (*p*-value = 0.067) (**Figure 4F**) indicating that the relative abundance pDCs are higher under inflammatory conditions, thereby supporting our hypothesis.

**Figure 4.**
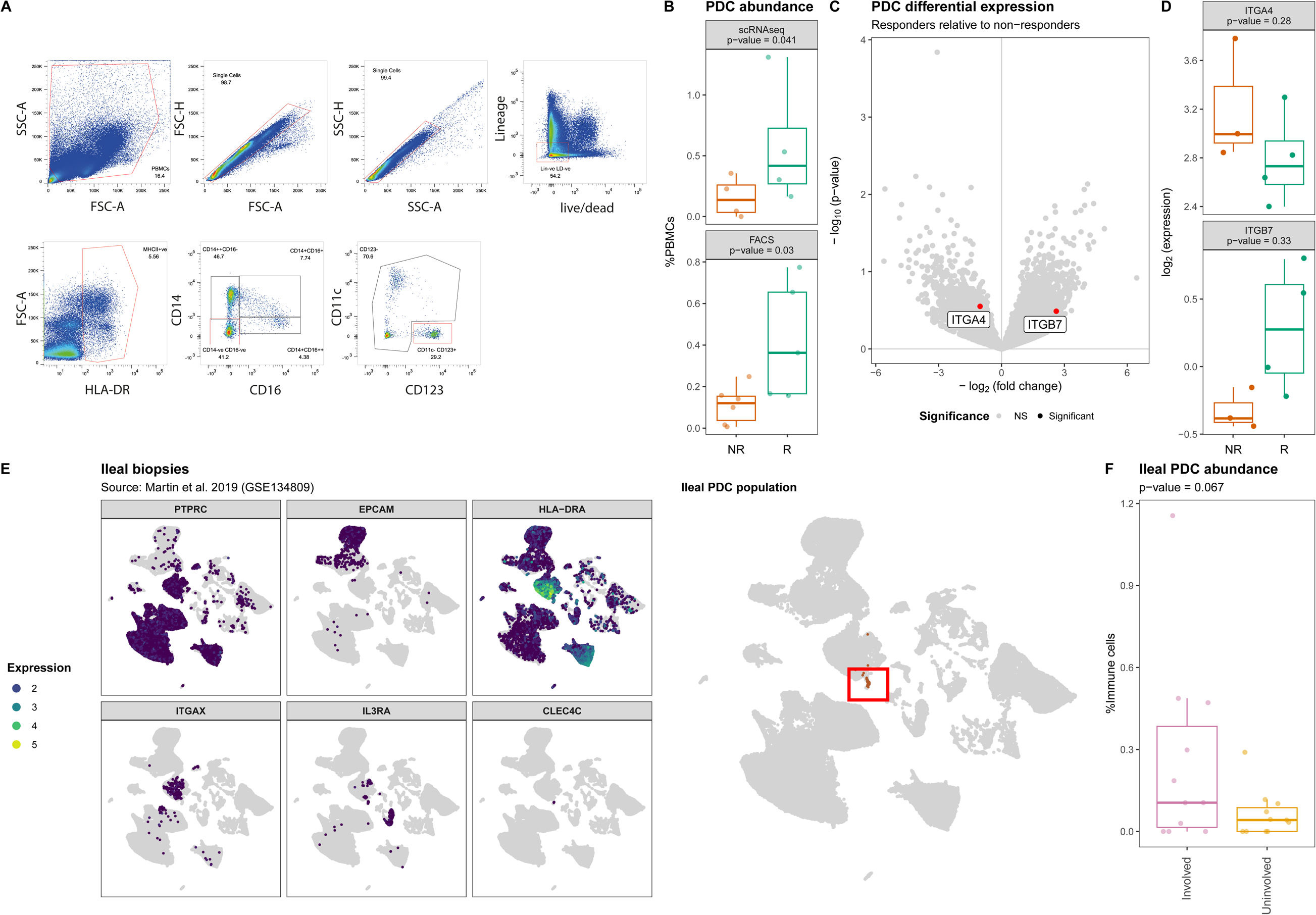
Lower abundance of plasmacytoid dendritic cells in PBMCs of non-responding patients. (A) Flow cytometry strategy used to identify and quantify the HLA-DR^+^CD14^-^CD16^-^CD11c^-^ CD123^+^ pDCs where red boxes indicate selected events. (B) Boxplot visualizations of the pDC abundances relative to all measured PBMCs annotated with the *p*-value obtained through t-test. (C) Volcanoplot comparing pDCs from responders with non-responders where the X-axis represents the log_2_(fold-change) and the Y-axis the –log_10_(*p*-value). Highlighted in red are *ITGA4* and *ITGB7*. (D) Boxplot visualizations of the *ITGA4* and *ITGB7* expression in pDCs showing visible but no statistical significant differences between responders and non-responders. *P*-values were calculating using the t-test implementation in speckle::propeller. (E) UMAP visualization of GSE134809 showing (left) the identification strategy of the PTPRC[CD45]^+^EPCAM^-^HLA-DRA^+^ITGAX[CD11c]^-^ IL3RA[CD123]^+^CLEC4C[BDCA2]^+^ pDCs, (right) as highlighted by the red box. (F) Boxplot visualization of the ileal pDC abundance relative to all immune cells colored by whether they originate from lesions (involved) or outside a lesion (uninvolved) shows that lesional pDCs are more abundant than non-lesional pDCs.

### Classical monocytes from VDZ non-responders present an altered transcriptome

UMAP visualization of the monocytes indicated response-associated clustering (**Figure 5A**), which was most visible for the classical monocytes, suggesting transcriptome-wide differences. Differential expression analysis of the classical monocytes identified 30 statistically significant differentially expressed genes (DEGs) (**Figure 5B** and **Supplemental Table 6**). Notably, responders presented higher expression of several monocyte/macrophage-function related genes including genes encoding cytokines (CXCL2 (21, 22), CCL3 (23–25), CCL4 (26, 27)), mediators of host defense signaling (RIPK2 (28)), and macrophage scavenging receptor (MSR1 (29)), typically observed in M2-like macrophages. By contrast, expression of genes encoding complement factor D (*CFD*) and negative regulator of NFκB signaling pathway (*VSTM1*) (30) was higher among non-responders (**Figure 5C**). We were able to confirm differential expression for *CFD* and *MSR1* through bulk RNA-sequencing on sorted classical monocytes (**Figure 5D** and **Supplemental Table 7**). Specifically interrogating *ITGA4* and *ITGB7* indicated neither significant nor visible differences in the gene expression thereof (**Figure 5E**). Gene set enrichment analysis against the KEGG database identified general lower expression of the cytokine-cytokine receptor signaling pathway among non-responders (**Figure 5F-G** and **Supplemental Table 8**). We were therefore interested in identifying which other PBMCs were the sender/receiver to the differentially expressed cytokines produced by the classical monocytes. Among classical monocytes derived from responders, we observed a significantly higher expression of vascular endothelial growth factor (VEGF). Notably, VEGF receptor 1-encoding *FLT1* was found to be higher in both the CD4T naïve as well as the classical monocytes (**Figure 5H**), suggesting a more wound-healing phenotype in classical monocytes obtained from responders.

**Figure 5.**
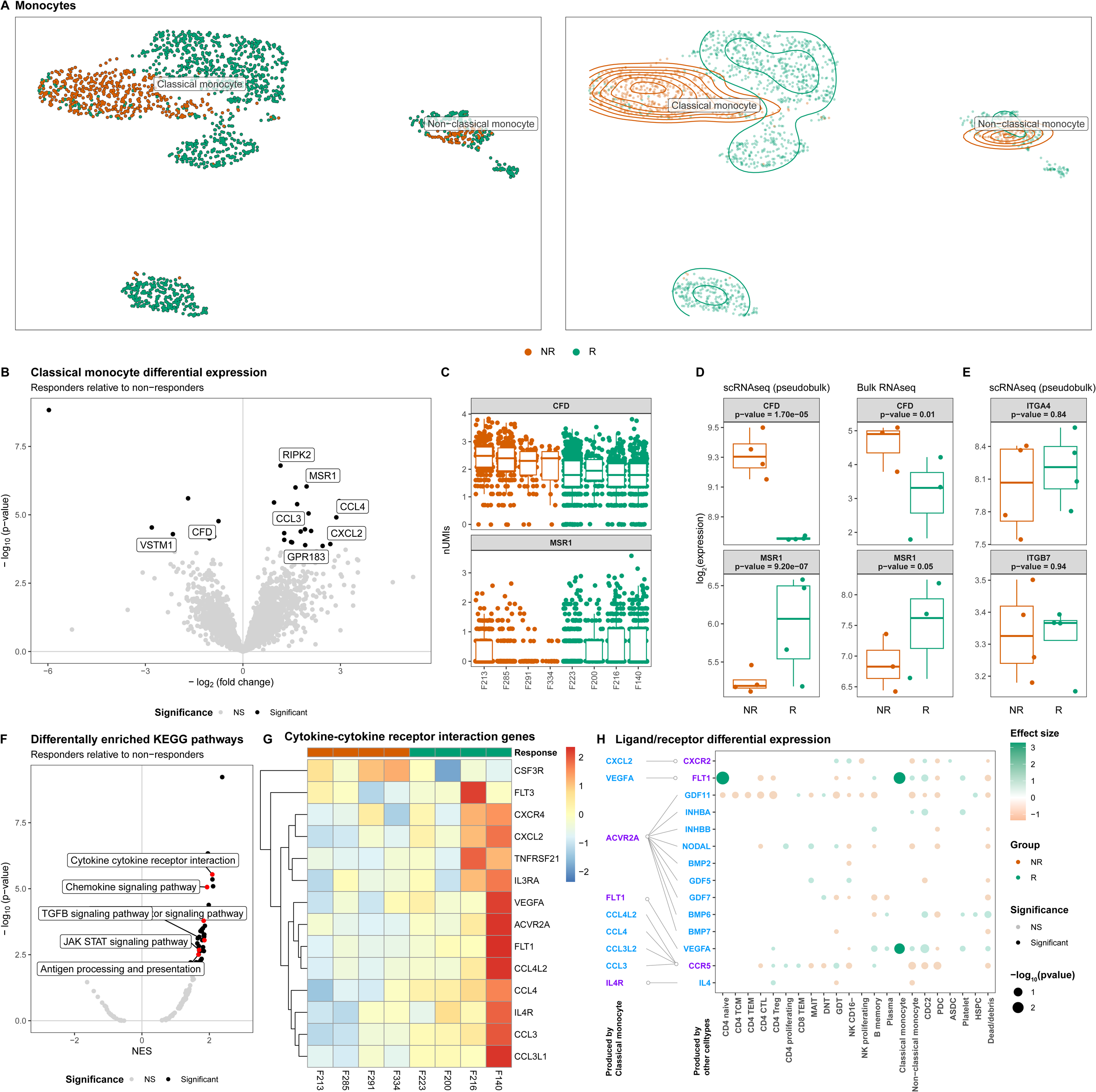
Classical monocytes from non-responding patients present lower expression of cytokine-cytokine signaling. (A) UMAP visualization of the monocytes colored by colored by response as dots (left) and a density plot (right) shows distinct clustering by response for the classical monocytes in particular. (B) Volcanoplot comparing classical monocytes from responders with non-responders where the X-axis represents the log_2_(fold change) and the Y-axis the –log_10_(*p*-value). Statistically significant differences (*p*-value_BH-adjusted_<0.05) are depicted in black. (C) Boxplot visualizations of *CFD* and *MSR1* expression in classical monocytes colored by response and grouped by patient where each dot represents an individual cell. Boxplot visualizations of (D) *CFD* and *MSR1*, and (E) *ITGA4* and *ITGB7* where the Y-axis represents gene log-transformed (left) normalized pseudobulk expression from our scRNAseq experiment and (right) normalized expression obtained through bulk RNAseq analysis on classical monocytes colored and grouped by response. *P*-values were obtained through Wald test as implemented in DESeq2. (F) Volcanoplot comparing classical monocytes from responders with non-responders where the X-axis represents the normalized enrichment score (NES) and the Y-axis the –log_10_(*p*-value). Statistically significant differences (*p*-value_BH-adjusted_<0.05) are depicted in black with pathways of interest highlighted in red. (G) Heatmap visualization genes belonging to the cytokine-cytokine receptor interaction pathway. Values represent the pseudobulk expression per sample, where color is proportion to the level of expression. (H) Dotplot visualization of a receptor/ligand interaction analysis of the differentially expressed cytokines representing ligands and receptors are colored in blue and purple, respectively. Depicted in the dotplot are the binding partners of the cytokines found to be differentially expressed by the classical monocytes for each cell type found in PBMCs. Size of the dots represents statistical significance, transparency the significance threshold, and color whether the gene is upregulated in either responders (green) or non-responders (orange). We find that VEGFA receptor FLT1 is significantly higher expressed among the CD4 naïve and classical monocytes of responders.

## Discussion

We demonstrate that VDZ-treated CD patients differ in cellular composition and intrinsic cellular behavior when comparing responders with non-responders. Interrogating the T cell compartment suggested a higher abundance of MAIT and CD8 TCM among non-responders. Notably, both T cells from non-responders appeared to be transcriptionally primed for NFκB signaling, a signaling pathway typically reserved for inflammation, by downregulating inhibitors thereof. The priming of the NFκB pathway would match the non-responding phenotype, where inflammation is still present despite treatment. While our results confirm that response to VDZ affects the T cell compartment, we also find response-associated differences within the myeloid compartment. We observed that the circulatory pDCs were significantly less abundant amongst non-responders. The pDC population represents a unique cell type whose ontogeny and lineage affiliation remain under debate due to its similarity to both myeloid and lymphoid lineages (31–35). Previously, pDCs were called “natural interferon producing cell” as they can produce large amounts of type I interferons (IFN), which typically occurs in response to viruses. This in turn activates NK and B cells (36–39), thereby bridging the innate and adaptive immune system. Remarkably, pDCs constitute only 0.4% of all measured cells when looking at all measured PBMCs in our scRNAseq experiment and only 0.12% of the immune compartment when interrogating ileal tissue from Martin *et al.* (20). Despite the rarity of the pDC population, they have been implicated in multiple immune-mediated inflammatory disorders (IMIDs) (35) evidenced by an ongoing phase II clinical trial testing the efficacy of litifilimab, a monoclonal antibody against pDC-specific blood dendritic cell antigen 2 (BDCA2), in systemic (40) and cutaneous (41) lupus erythematosus to dampen type I IFN production (42, 43). Despite the established role of pDCs in other IMIDs, their association with CD or IBD as a whole is less well documented. Previous studies have indicated that the circulatory pDC population is significantly decreased in IBD patients with active disease (44), with subsequent research by the same authors showing increased infiltration into the colonic mucosa and mesenteric lymph node (MLN) (45). This largely corroborates our own observations, as samples were obtained during treatment and the difference between responders and non-responders is by definition a difference in inflammation. However, controversy exists on what role pDCs play in the pathogenesis of IBD as experiments have yielded conflicting results. It has been reported that pDCs can aggravate (46), protect (47, 48), or are dispensable in the development of experimental IBD (49). Accordingly, it remains unclear how pDCs might play a role in responsiveness towards VDZ.

Significant differences in expression were observed for the circulating classical monocytes, which presented a more scavenger-like, wound-healing phenotype amongst responders. By contrast, classical monocytes from non-responders appeared to present higher expression of *CFD*, a gene involved in the alternative complement pathway. Complement factor D cleaves factor B forming Bb, thereby activating the complement cascade (50). The alternative complement pathway is an important component of the innate immune response where it is typically used as first line defense against microbes. Our results would imply that the classical monocytes from non-responders are primed at activating the alternative complement pathway. Such monocytes could potentially be recruited into the intestinal compartment, where they would differentiate into macrophages. Intestinal inflammatory macrophages are one of the few macrophages that are purported to be supplemented by the circulating monocyte population during inflammatory episodes (51–53). We show that monocytes indeed express genes encoding components that form integrin α_4_β_7_, corroborating earlier observations by Schleier *et al.* who showed that monocytes present functional integrin α_4_β_7_ on their surface, with VDZ abrogating their interactions with MAdCAM-1 *in vitro* (14). While our observations do not indicate any difference in expression of either *ITGA4* or *ITGB7*, we note that the classical monocytes, alongside all other myeloid cells, were less abundant among non-responders, which we hypothesize is due to their recruitment out of circulation and into the intestinal compartment.

Taken together, it is evident that response to VDZ during treatment manifests itself in various circulating cell types, presenting differences in both abundance as well as expression. While the current study provides novel insights into the diagnostic capabilities single-cell transcriptomics for elucidating response to VDZ, we acknowledge the shortcomings of this study in terms of the limited sample size as well as the largely associative nature of the observations. More importantly, patient-samples taken during treatment do not hold prognostic value in predicting response to therapy. Future experiments would need to be conducted to validate the differences observed at the level of single-cell transcriptomics in a larger independent cohort to fully understand its utility as biomarker. Furthermore, to disentangle inflammation from VDZ-treatment, samples would need to be included prior to the start of treatment. Moreover, it is imperative to compare our observations with CD patient cohorts treated with other inflammatory-reducing medication to understand which observations VDZ-specific and which observations are inflammation-associated. Such an approach would not only allow for the identification of prognostic biomarkers for VDZ response, but also provide potential targets that might be involved in the manifestation of drug non-response. Taken together, we demonstrate that patients on VDZ treatment present differences in the cellular heterogeneity of PBMCs. Further confirmatory studies are necessary to understand the full potential of the observed differences.

## Methods

### Cohort assembly and sample collection

Patients included were obtained from the EPIC-CD study, which is a multi-center consortium with the goal of identifying prognostic biomarkers at the level of peripheral blood (PBL) DNA methylation capable of predicting response to adalimumab, infliximab, vedolizumab, and ustekinumab prior to treatment in CD patients (54). For the current study, 8 VDZ-treated CD patients (4 responders and 4 non-responders) were sampled for PBL at a median of 41 weeks (interquartile range 31-70) into treatment during routine care at the AmsterdamUMC hospital, location AMC, Amsterdam, Netherlands between 2018 and 2019 (**Table 1**). Adult CD patients were included if they were presented with active ulcerative disease as defined by a simple endoscopic score CD (SES-CD) >3. Response was assessed at approximately 41 weeks based on a reduction relative to the baseline measurement in endoscopic (ΔSES-CD≥50%) in combination with either clinical-(ΔHarvey Bradshaw Index (HBI)≥3 or HBI≤4) and/or biochemical (ΔC-reactive protein (CRP)≥50% or CRP≤5 g/mL or Δfecal calprotectin (FCP)≥50% or FCP≤250 µg/g) criteria in the absence of systemic corticosteroids. Immediately after collecting peripheral blood, peripheral blood mononuclear cells (PBMC) were isolated by means of Ficoll (GE Healthcare) separation and IMDM medium (Gibco) supplemented with 10% DMSO and 50% FBS (Serana). Isolated PBMCs were stored overnight at −80 C in Mr. Frosty freezing containers (Thermo) where after they were transferred to liquid nitrogen until cohort completion.

### General bioinformatic data analyses

Data was imported and analyzed using the R statistical environment (v4.2) (55) using several packages obtained from the Bioconductor (v3.16) (56) repository. The analytical workflow was orchestrated by Snakemake (v7.14.0) (57). Visualizations were created using the *tidyverse* (v1.3.1) (58), *ggplot2* (v3.4.2) (59), *ggrastr* (v1.0.1), *ggrepel* (v0.9.3), *cowplot* (v1.1.1), *viridis* (v0.6.3) (60), *pheatmap* (v1.0.12).

### Single-cell RNA-sequencing analysis

Samples were removed from the cryostat and thawed on ice. Thawed PBMCs were washed and then labelled using the BioLegend TotalSeq-B cell hashtag oligo (HTO) antibodies for multiplexing purposes per manufacturer’s protocol at 1 U per 1 million cells (61). An aliquot of the tagged PBMCs was assessed for viability using the Countess II FL Automated Cell Counter indicating that over 80% of the cells were viable. The resulting oligo-tagged cell suspensions were then mixed and distributed across 6 GEM-wells to be loaded onto the Chromium controller (10X Genomics) using 10X chemistry v3. Per well, 10,000 cells were loaded for a= targeted recovery rate of up to 6,000 cells. Single-cell barcoded partitions were prepared using 10X chemistry v3 where after separate sequencing libraries were prepared for HTOs and the actual mRNA after size-selection. Libraries were sequenced on the Illumina HiSeq4000 in a 150 bp paired-ended fashion at the Core Facility Genomics, Amsterdam UMC. The mRNA libraries were sequenced on 150M reads per GEM-well, whereas the HTO libraries were sequenced to a depth of 50M reads per GEM-well.

Resulting raw reads were aligned and unique molecular identifier (UMI) count matrices were generated using Cellranger (v7.0.0) (10X Genomics). Subsequent import, sample-wise demultiplexing, processing, and analysis was done in Seurat (v4.3.0) (62). Cells that were identified as multiplets, based on the presence of an equal number of different HTOs, or that did obtain sufficient HTO signal were removed as they could not be assigned to a unique donor. Subsequent quality control included identifying dead cells based on mitochondrial read content (>75%) and a low number of unique genes, which were annotated accordingly (63). UMI counts were normalized using SCTransform (64) using default. Cells were subsequently annotated by mapping our data onto a reference PBMC CITE-seq experiment of 162,000 annotated cells using a weighted nearest neighbor approach (65, 66). A subsequent manual curation using canonical markers confirmed the identity of the different cell types at cluster level. A dead and debris cluster was identified as cells with a low number of unique genes (<500 unique genes) and a high percentage mitochondrial reads (>80%), multiplet were identified on hashtags derived from multiple different donors (inter-donor multiplets), and/or a high number of unique genes (>2000 unique genes) in combination with multiple celltype-specific markers (mixed-cell multiplets). Proliferating cells were identified based on their high expression of proliferation marker *MKI67* (67). T cells were identified based on the expression of *CD3D*, *CD2*, *CD7*, and *IL7R*. Natural killer (NK) cells were identified based on the expression of *CD2*, *CD7*, *GNLY* and *NKG7*, while lacking *CD3D*, B-cells were identified based on the expression of *MS4A1* and *BANK1* positive. Monocytes were identified based on the expression of *CST3* and *CD14* or *FCGR3A*. Conventional dendritic cells (cDC) were identified on expression of *CD1C*, *CST3*, *FCER1A* and *HLA-DRA*. Smaller cell populations not belonging to the lineages were identified as well, namely the thrombocytes (*CST3* and *PPBP* positive), hematopoietic stem and progenitor cells (HSPCs; *CD34* positive), and erythroblasts (*HBA1*, *HBA2*, *HBQ1*, and *HBB* positive) (68).

### Mass cytometry by time-of-flight

Concurrent with the single-cell RNA-sequencing analyses, mass cytometry by time of flight (CyTOF) was performed on a separate aliquot of the PBMC samples. Here, we measured the cell-surface expression of 37 proteins with a particular focus on the T cell lineage. An overview of all the antibodies used and their clones can be found in **Supplemental Table 1**. Cryopreserved PBMCs were thawed, washed with PBS, and resuspended in RPMI medium. Cellular viability was assessed through live/dead staining using 5μM Cisplatin in PBS at room temperature. Cisplatin signal was quenched by washing with Cell Staining Buffer (CSB; Fluidigm) after 5 minutes and washed away. As several targets in the panel are known to lose their binding specificity after PFA fixation, the corresponding antibodies were incubated in the presence of Human TruStain FcX™ Fc Receptor Blocking Solution (BioLegend) at room temperature for 30 minutes. After washing, cells were fixed with 1.6% PFA and labeled using the Cell-ID 20-Plex Pd Barcoding Kit (Fluidigm) for multiplexing purposes per manufacturer’s protocol. Pooled cells were then stained for remaining cell-surface targets. Antibody concentrations were optimized for staining 3M cells per 100 μL of CSB for 30 minutes at room temperature. For intracellular staining, cells were washed and incubated with antibodies for intracellular markers (CES1 and CTLA4). CES1 lacked a metal reporter but was the only rabbit anti-human antibody, so a goat anti-rabbit antibody coupled to ^175^Lu was used as a secondary staining for CES1. After washing with CSB, antibodies were again fixed with 1.6% PFA, washed and incubated overnight with ^191/193^Ir DNA intercalator (1:4000) diluted in Fix-and-Perm Buffer (Fluidigm). Cells were subsequently washed before data acquisition was performed on the CyTOF3-Helios (Fluidigm). After data acquisition, raw .FCS files were imported in R. Expression values were arcsinh-transformed with cofactor 5. Signal intensities and sample acquisition rates were reviewed for stability over time and events gated based on the condition that the flow was stable, excluding calibration beads, and within the 90th percentile of all Gaussian parameters. Resulting singlets were selected for CD45^+^ signal. Cells were clustered in an unsupervised manner using the FlowSOM-package, where initial SOM-clustering was set to 300 clusters, using markers listed in **Supplementary Table S1**. The 300 clusters were subsequently manually metaclustered according to their phenotypic lineages, whereafter cells were annotated. UMAP dimensionality reduction was performed using the *uwot* (v0.1.14). A total of 16,000 cells were randomly selected without replacement to approximately match the number of cells identified through scRNAseq, making the observations more comparable.

### Flow cytometry of the plasmacytoid dendritic cells

In addition to using an aliquot of PBMCs of the same patients analyzed for scRNAseq and CyTOF, an additional two patients (1 responder and 1 non-responder) were included in the flow cytometry analyses. Upon thawing, PBMCs were washed in PBS and stained for a live/dead cell viability marker (LifeScience, Amsterdam, the Netherlands). Cells were subsequently stained for surface markers in FACS buffer (0.5% BSA, 0.01% NaN3 in PBS) using the following antibodies: CD11c-PerCP Cy5.5 (clone: S-HCL-3, BioLegend), HLA-DR-Alexa Fluor 700 (clone: LN3, eBioscience), CD123-FITC (clone: 6H6, BioLegend), CD1C-PE-Cy7 (clone: L161, BioLegend), pan-lineage (CD3/CD19/CD20/CD56)-APC (clones: UCHT1;HIB19;2H7;5.1H11, BioLegend), CD14-BD Horizon V500 (clone: M5E2, Becton Dickinson) and CD16-PE (clone: 3G8, Becton Dickinson). Acquisition was performed on the BD LSR Fortessa™. Doublets were excluded and live single cells identified using the forward scatter height (FSC-H) versus the forward scatter area (FSC-A) and the side scatter height (SSC-H) versus side scatter area (SSC-A). Live cells were identified using the live/dead marker. Classical monocytes were defined as (T/B/NK) Lin^-^HLA−DR^+^CD14^++^CD16^-^, intermediate monocytes as Lin^-^HLA−DR^+^CD14^++^CD16^+^, and non-classical monocytes as Lin^-^ HLA-DR^+^CD14^+^CD16^+^. Conventional dendritic cells (cDCs) were defined as (T/B/NK) Lin^-^ HLA-DR^+^CD11c^+^CD1c^+^ and plasmacytoid DCs (pDCs) as Lin^-^HLA-DR^+^CD11c^-^CD123^+^. Fluorescence minus one (FMO) was used for gating and median fluorescence intensity was determined to quantify cell surface expression. An overview of all antibodies and their clones used can be found in **Supplemental Table 2**.

### RNA-sequencing of the classical monocytes

Akin to the flow cytometric analyses, PBMCs were washed in PBS and stained for a live/dead cell viability marker (LifeScience, Amsterdam, the Netherlands) alongside the antibodies mentioned above. Cell sorting was conducted on the SH800 Cell Sorter (Sony). Classical, intermediate, and non-classical monocytes were identified as (T/B/NK)Lin^-^HLA-DR^+^CD14^++^CD16^-^, HLA-DR^+^CD14^++^CD16^+^, and HLA-DR^+^CD14^+^CD16^+^, respectively. The classical monocytes were sorted out and were subsequently processed for RNA sequencing. Due to low input material, classical monocytes mRNA was converted into cDNA using the Ovation RNA-seq System V2 kit (NuGEN; Agilent, Santa Clara, United States), whereupon sequencing libraries were prepared using the Ovation Ultralow System V2 kit (NuGEN; Agilent, Santa Clara, United States) and thereafter sequenced in a 150 bp paired-ended fashion on the Illumina NovaSeq6000 to a depth of 40 million reads at the Amsterdam UMC Core Facility Genomics. Quality control of the raw reads was done using FastQC (v0.11.8) (69) and MultiQC (v1.0) (70). Raw reads were aligned to the human genome (GRCh38) using STAR (v2.7.0) and annotated using the Ensembl (v95) annotation (71). Post-alignment processing was performed through SAMtools (v1.9) (72), after which reads were counted using the featureCounts function found in the Subread package (v1.6.3) (73). Differential expression (DE) analysis was performed using *DESeq2* (v1.36.0) (74).

### Statistics

Differential abundance analyses were conducted by comparing the proportions using a t-test as implemented in the *speckle* (v0.99.7) (75) package where we omitted cell types that were represented by 10 cells or less. Differential expression analyses were conducted using the “pseudobulk” approach (76). Cells per sample per donor to account for cells coming from the same donor. The actual differential expression analyses were performed using the Wald test as implemented in the *DESeq2* (v1.36.0) (74) package. Subsequent gene set enrichment analyses were conducted using Wald statistic as input for *fgsea* (v1.22.0) (77) against the Kyoto Encyclopedia for Genes and Genomes (KEGG) gene sets (78).

### Study approval

All included patients provided informed consent and the sampling was in accordance with the institutional ethics committee (METC reference number: NL53989.018.15).

## Data Availability

Raw data produced are available online at the European Genome-phenome Archive. The single-cell RNA-sequencing data can be found under accession number EGAS00001007328. The bulk RNA-sequencing data on classical monocytes can be found under accession number EGAS00001007361. All scripts can be found at https://github.com/ND91/HGPRJ0000008_EPICCD_anti_a4b7_sc

## Data availability

The data that support the findings of this study are available under controlled access. The raw sequencing data can be found at the European Genome-phenome Archive EGAS00001007328 and EGAS00001007361 for the single-cell RNA-sequencing data and the bulk RNA-sequencing on classical monocytes, respectively. All scripts can be found at https://github.com/ND91/HGPRJ0000008_EPICCD_anti_a4b7_sc.

## Author contributions

ALY: Study design, conducting experiments, acquiring data, analyzing data, writing the manuscript.

IH: Study design, conducting experiments, acquiring data, patient material acquisition, analyzing data, writing the manuscript.

VJ: Study design, acquiring data, patient material acquisition, writing the manuscript.

AE: Conducting experiments, acquiring data, analyzing data.

MG: Conducting experiments.

EL: Analyzing data

JV: Conducting experiments, acquiring data, analyzing data.

CV: Conducting experiments.

IAvdB: Conducting experiments.

MM: Supervision.

MJ: Conducting experiments, acquiring data.

SK: Conducting experiments, acquiring data.

AA: Analyzing data.

JS: Supervision

GD: Patient material acquisition, supervision.

WdJ: Study design, supervision, funding acquisition.

PH: Study design, supervision, funding acquisition.

All authors have read and agreed to the published version of the manuscript.

## Acknowledgements

This work was partly funded by the Helmsley Foundation as well as local funds from the Department of Human Genetics and the Tytgat Institute for Liver and Intestinal Research, AmsterdamUMC University of Amsterdam, Amsterdam, Netherlands. The study sponsors had no role in study, design, collection, analysis, or interpretation of data.

We are thankful to all the participants included in the EPIC-CD study as well as the Core Facility Microscopy and Cytometry and the Core Facility Genomics located at the AmsterdamUMC for their expert assistance in the single-cell RNA-sequencing, mass cytometry, and flow cytometry experiments and analyses.

## Supplementary Tables

**Supplemental Table 1. Marker panel mass cytometry**. The cell-surface exposed markers assayed in the mass cytometry experiment annotated by the metal, target protein, alternative names, uniport identifier, and notes.

**Supplemental Table 2. Marker panel flow cytometry**. The cell-surface exposed markers assayed in the flow cytometry experiment alongside the antibody and clone.

**Supplemental Table 3. PBMC differential abundance analysis.** Results of the differential abundance analysis on the major lineages as conducted using the propeller function in speckle. Columns represent the cell type, the mean proportion for all samples (“BaselineProp.Freq”), non-responders only (“PropMean.Non.responder”), and responders only (“PropMean.Responder”), the ratio responder/non-responder (“PropRatio”), the associated T statistic (“Tstatistic”) as well as the *p*-values (“P.Value” and “FDR”). Tabs separate the results from scRNAseq and CyTOF analyses.

**Supplemental Table 4. T cell scRNAseq pseudobulk differential gene expression analysis.** Results of the differential expression analysis performed on PDCs using DESeq2. Columns represent the gene, the average expression (“baseMean”), the log_2_(fold change) (“log2FoldChange”), the associated standard error (“lfcSE”), the Wald statistic (“stat”) as well as the *p*-values (“pvalue” and “padj”). Each tab represents a different T cell subset.

**Supplemental Table 5. pDC scRNAseq pseudobulk differential gene expression analysis.** Results of the differential expression analysis performed on PDCs using DESeq2. Columns represent the gene, the average expression (“baseMean”), the log_2_(fold change) (“log2FoldChange”), the associated standard error (“lfcSE”), the Wald statistic (“stat”) as well as the *p*-values (“pvalue” and “padj”).

**Supplemental Table 6. Classical monocytes scRNAseq pseudobulk differential gene expression analysis.** Results of the differential expression analysis performed on classical monocytes using DESeq2. Columns represent the gene, the average expression (“baseMean”), the log_2_(fold change) (“log2FoldChange”), the associated standard error (“lfcSE”), the Wald statistic (“stat”) as well as the *p*-values (“pvalue” and “padj”).

**Supplemental Table 7. Classical monocytes bulk RNAseq gene expression analysis.** Results of the differential expression analysis performed on classical monocytes using DESeq2. Columns represent the gene and Ensembl ID, the average expression (“baseMean”), the log_2_(fold change) (“log2FoldChange”), the associated standard error (“lfcSE”), the Wald statistic (“stat”) as well as the *p*-values (“pvalue” and “padj”).

**Supplemental Table 8. Classical monocytes scRNAseq pseudobulk KEGG gene set enrichment analysis**. Gene set enrichment analysis as performed by fgsea. Columns represent the gene set (“Pathway”), the *p-*values (“pvalue” and “FDR”), the log_2_ standard error (“log2err”), the enrichment score (“ES”), the normalized enrichment score (“NES”), and the total number of genes in the gene set (“size”).

